# A prospective study of pre-operative risk factors associated with extended length of stay in patients attending a pre-operative clinic in South Africa

**DOI:** 10.1101/2024.10.13.24315313

**Authors:** Kuven Naidu, Nabeela Kajee, Jayseelan Naidu, Bilaal Wadee

## Abstract

**Background:** Preoperative assessment clinics play a critical role in identifying, evaluating, and mitigating perioperative risks. Despite global data highlighting the importance of preoperative risk factors on surgical outcomes, there remains limited information on their impact on postoperative length of stay in South African contexts.

**Objectives:** This study aimed to describe the demographic and clinical profiles of patients referred to a preoperative clinic in Johannesburg, South Africa, and to examine factors associated with postoperative length of stay (LOS).

**Methods:** This was a prospective cohort study conducted between 2021 and 2022 at a private clinic. Patients aged ≥18 years undergoing non-cardiac surgery were included. Data on demographics, comorbidities, surgical procedures, and clinical outcomes were collected. Statistical analysis was performed to assess relationships between preoperative risk factors, including ASA grading, Revised Cardiac Risk Index (RCRI), eGFR, Diabetes Mellitus, age, obesity and LOS.

**Results:** A total of 214 patients were assessed, of which 75.7% were female, with a median age of 62.5 years. Common comorbidities included hypertension (59.3%) and obesity (55%). The median LOS was 3.5 days, with 47.2% of patients staying more than 3 days postoperatively. Knee (33.2%) and hip surgeries (21%) were the most common procedures. A significant association was found between longer LOS and RCRI score ≥1 (p=0.007), as well as renal dysfunction in knee surgery patients (p=0.027) and age in patients undergoing hip surgery (p=0.049). There was no significant association between ASA grade, presence of diabetes mellitus, eGFR, age or obesity.

**Conclusions:** Increased RCRI scores, renal dysfunction, and age were associated with prolonged hospital stay. Findings highlight the need for targeted interventions in preoperative care to reduce LOS, particularly for high-risk patients. Further research is needed to validate these results across broader healthcare settings and to establish appropriate protocols for South African patients undergoing pre-operative assessment.

## Introduction

Preoperative assessment is an integral aspect of surgical care, offering a unique opportunity to identify, evaluate, and mitigate perioperative risks. The effectiveness of a preoperative clinic can dramatically affect patient outcomes and healthcare resource allocation. With over 234 million major non-cardiac surgeries performed globally each year, perioperative complications, including major adverse cardiac events (MACE), significantly contribute to patient morbidity, mortality and prolonged length of stay^(1)^. Preoperative clinics play a critical role in identifying medical issues, either new or old, that may impact on a patients surgical journey^(2)^. It has been established that preoperative risk factors are a significant predictor of increasing hospital costs more so than actual procedure complications themselves ^(3)^.

Preoperative clinics have been shown to decrease lxsength of stay ^(4)^ ^(5)^ and play important roles in decreasing unnecessary and unwarranted consultations ^(6)^. Patients who attended a preoperative clinic were also less likely to have their procedures cancelled on the day of admission for the procedure by an anaesthetist ^(5)^. This can be a source of frustration for the patient, the doctor and the facility. Prolonged length of stay increases the cost per admission. A South African study has estimated the cost per day in ICU to be approximately R22870 per day in the public sector^(7)^. A day in the surgical general ward would cost approximately R4184 per day^(8)^. It will therefore be important to identify any possible causes beforehand that may affect length of stay in hospital post procedure. The revised cardiac risk index (RCRI) score can identify patients undergoing noncardiac surgery in whom complications are more likely to occur ^(9)^. A modified risk score of greater than or equal to 3 has also been shown to have increased non-cardiac morbidity as well as a prolonged hospital stay after elective orthopaedic procedures ^(10)^. However, there remains limited South African data on the value of using an RCRI as a risk factor for prolonged stay post-surgery. The American Society of Anesthesiologists (ASA) Grading system (I-VI) is used to determine the health of a patient before undergoing a procedure ^(11)^. A prolonged length of stay has also been noted with an increase from ASA Class I to ASA Class IV ^(12)^. Co-morbidities may also play an important role in prolonging length of stay post procedure. Age ^(13)^, Diabetes Mellitus ^(14)^, hypertension ^(15)^, obesity^(16)^, anaemia ^(17)^ and impaired renal function ^(18)^ have all been noted to play a role in extending length of stay post operatively.

The objectives of this study were to describe the demographic and clinical profile of patients referred for a pre-operative assessment to a private clinic in the East Rand of Johannesburg. We looked at the post operative length of stay in this cohort of South African patients and possible associated risk factors. This will add to the currently limited data available for South African centres hopefully allowing for further insight into risk stratification and appropriate resource planning.

## Materials and Methods

### Ethics Statement

Ethics was obtained from the National Health Research Ethics Committee of Life Healthcare Group: REC 251015-048 (approval number)

Written consent was obtained from each participant prior to any information being recorded.

### Study design, study setting and study population

This was a prospective cohort study based on record review. The cohort comprised of patients enrolled at the pre-operative clinic in a private practice in Benoni, Johannesburg between 25 Jan 2021 and 15 June 2022.

Inclusion criteria were any patient over the age of 18years referred for non-cardiac surgery and who were willing and able to give consent.

Exclusion criteria comprised of any patient under the age of 18 years and those unwilling to provide consent.

Data sources were the pre-operative assessment notes and length of stay was provided by the hospital to which the patients were admitted.

### Data collection and analysis

Data from the medical records was captured in a study specific Excel database. Data will be exported into Stata 18.0 [ Stata Corporation, College Station, USA] for analysis.

Demographics (age, sex, race, date of assessment) and clinical characteristics (comorbidities, hematological and biochemical results) of included participants will be described using categorical and continuous variables. Categorical variables will be described as frequencies and percentages. Continuous variables which are normally distributed will be described as means and standard deviations while variables which are not normally distributed will be described as medians with interquartile ranges. Appropriate graphs and charts will be used to visualize the data.

The length of hospital stay was determined as the number of days from admission to discharge and will be presented as a median and interquartile range overall.

## Results

A total of 214 patients were seen at the pre-operative clinic with a total of 213 cleared to proceed with surgery (Table 1).

**Table 1:** Demographic characteristics of patients seen at Pre-Op clinic (N=214)

| Variable | n(%) |
| --- | --- |
| Age (median , IQR) | 62.5 (53- 71) |
| Age (mean, S.D) | 61.5 ( 12.4) |
| Sex |  |
| Male n (%) | 52 (24.3) |
| Female n (%) | 162 (75.7) |
| Race (self identifying) |  |
| Black African n (%) | 22 (10.3) |
| White n (%) | 176 (82.2) |
| Indian n (%) | 16 (7.5) |
| Coloured n (%) | 0 (0) |
| Year of enrolment |  |
| 2021 n (%) | 154 (72.0) |
| 2022 n (%) | 60 (28.0) |

7 patients of the 213 cleared (3.3%) had their procedures surgeries postponed or cancelled prior to undergoing surgery.

The mean age of the patients was 61.4 years (median of 62.5years). The majority of the patients were female (75.7%).

The four most frequent co-morbidities were hypertension (n=127; 59.3%), dyslipidaemia (n=61; 28.5%), hypothyroidism (n=38; 17.8%) and Type 2 Diabetes Mellitus (n=30; 14%).(Figure 1)

**Figure 1:**
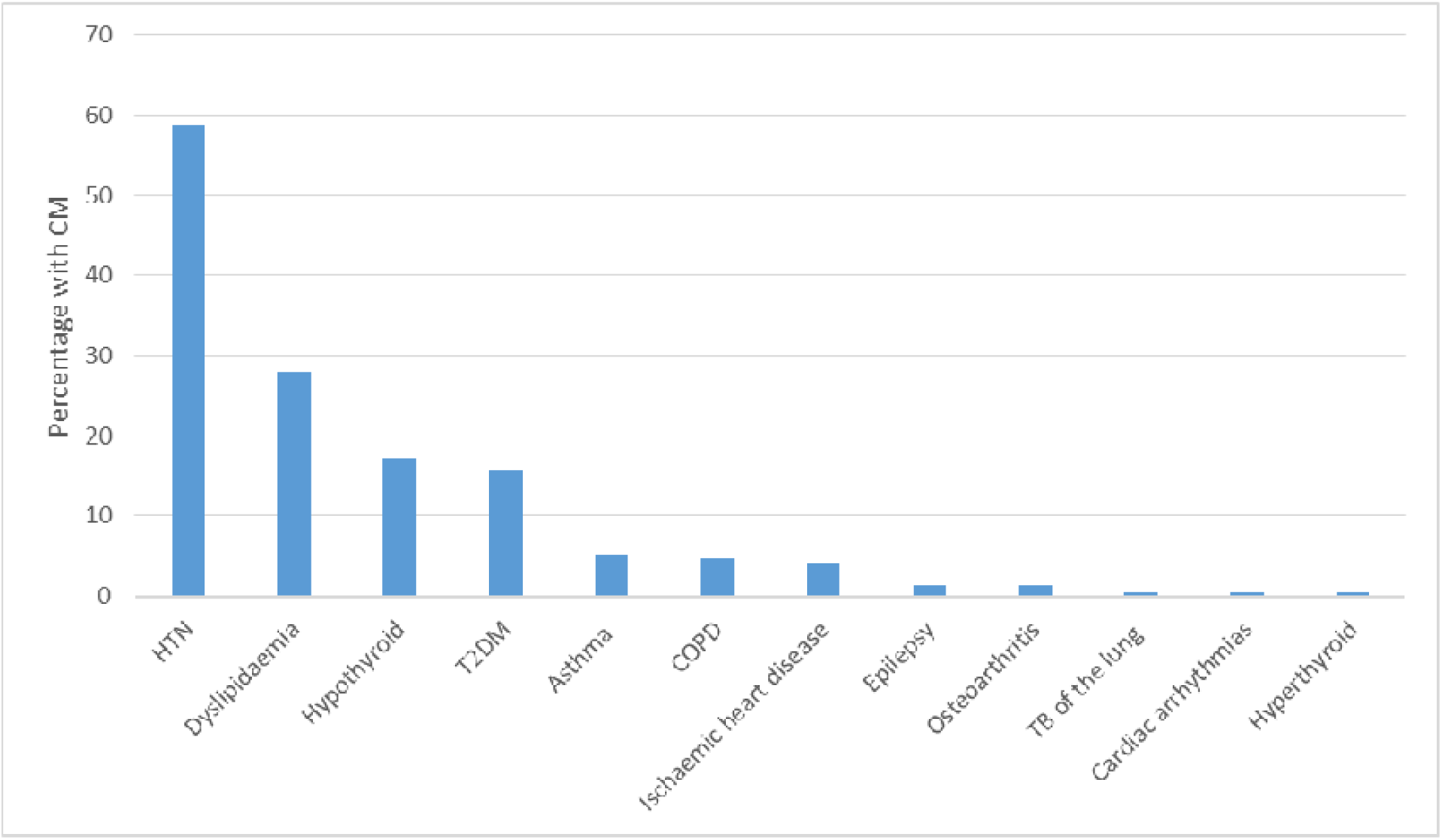
Frequency of comorbidities pre-operatively (N=214)

The majority of the patients were referred for elective knee surgery (n=71; 33.2%), hip surgery (n=45; 21%), hysterectomy or hysteroscopy and (n=43; 20%) and breast surgery (n=19; 8.9%).

There was Body Mass Index (BMI) data available in 211 of the patients with 55% being classified as obese (BMI >30kg/m^2^).

The median Blood pressure reading in the patients was 134/80.5 mmHg with 30.8% having a systolic blood pressure reading over 140mmHg and 11.2% having a diastolic reading above 90mmHg. (Table 2).

**Table 2:** Clinical profile of patients seen in the pre-operative clinic (N=214)

| Parameter | N | Median (IQR) | n(%) abnormal* |
| --- | --- | --- | --- |
| BMI (m/kg<sup>2</sup>) | 211 | 30.9 (27- 35.2) | 116 (55.0) |
| SBP (mmHg) | 214 | 134 (124- 143) | 66 (30.8) |
| DBP (mmHg) | 214 | 80.5 (74- 85) | 24 (11.2) |

\*BMI>30kg/m<sup>2</sup>; systolic blood pressure >140mmHg and diastolic blood pressure >90mmHg

The baseline biochemical profile of the patients is listed in Table 3.

**Table 3:** Biochemical profile of patients seen in the pre-operative clinic, N=214.

| Parameter | N | Median (IQR) | n(%) abnormal |
| --- | --- | --- | --- |
| Haemoglobin (g/dl) | 205 | 14.3 (13.3- 15.3) | 41 (20.0) |
| Haematocrit (L/L) | 201 | 40 (28 - 44) | 5 (2.5) |
| Sodium (mmol/l) | 203 | 140 (138- 141 ) | 19 (9.4) |
| Potassium (mmol/L) | 203 | 4.2 (3.9 - 4.4) | 15 (7.4) |
| Chloride (mmol/L) | 202 | 103 (101- 106) | 33 (16.3) |
| Urea (mmol/L) | 205 | 5.7 (4.4 - 6.8) | 39 (19.0) |
| Creatinine (umol/L) | 211 | 72 (62- 84) | 46 (21.8) |
| GFR (ml/min) | 204 | 83.06 (70.08 - 90.76) | 23 (11.3) |
| Random blood glucose (mmol/L) | 207 | 5.4 (5.0 - 6.2) | 12 (5.8) |

Normal values: Hb males 13.8-18.8 g/dL, Hb females 12.4-16.7 g/dl; HCT males 0.4-0.56 L/L, HCT females 0.35-0.49L/L females; Sodium 136-145 mmol/L; Potassium 3.5-5.1 mmol/L; Chloride 98-107 mmol/L; Urea 2.1- 7.1 mmol/L; Creatinine males 80-115umol/L, Creatinine females 53-97umol/L ; GFR >60ml/min ; Random blood glucose 4.0-11.1 mmol/L

The majority of the patients seen were graded as an ASA Grade 2 (81.2%) (Figure 3). 74.5% of patients had an RCRI score of 0 and 23.8% a score of 1. (Figure 4)

**Figure 3:**
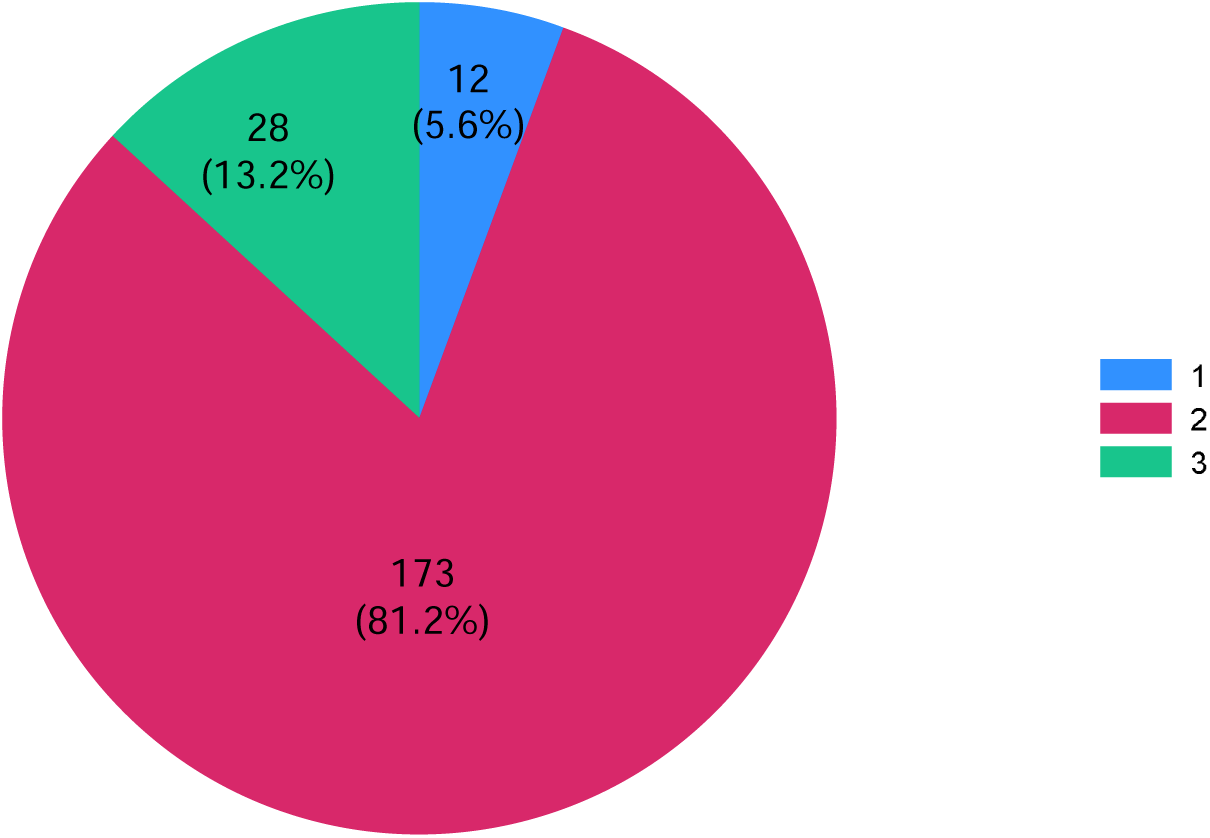
Distribution of ASA grading among patients seen in the pre-operative clinic (N=214)

**Figure 4:**
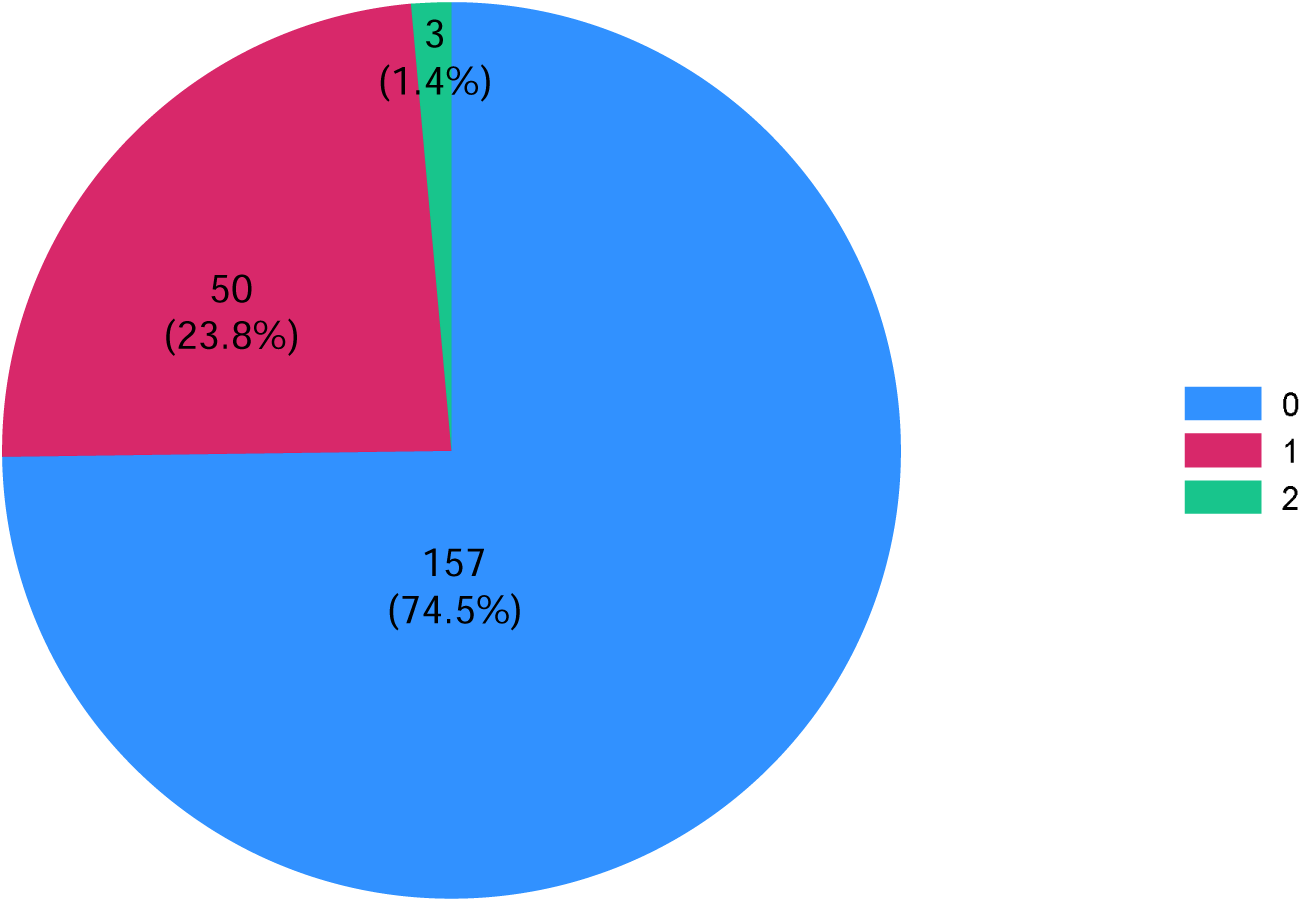
Distribution of RCRI points among patients seen in the pre-operative clinic, N=214.

Table 4 highlights the characteristics of patients who were admitted for longer than a day in hospital. A total of a 152 (71%) patients stayed for one day or longer. No significant differences were noted between the different surgical procedures other than patients undergoing uterine surgery had a larger proportion of patients (48.4%) with an RCRI score of 1 or 2.

**Table 4:** Characteristics of patients who stayed in hospital for one day or more by planned surgical procedure.

| Variable | Other (n=23) | Knee(n=47) | Hip (n=34) | Breast (n=17) | Uterine (n=31) | p-value |
| --- | --- | --- | --- | --- | --- | --- |
| Age (mean , sd) | 60.3 | 63.7 (10.3) | 63.2 | 67.5 | 51.9 | 0.267* |
|  | (14.1) |  | (10.7) | (10.4) | (10.3) |  |
| Male n (%) | 3<br>(13.0) | 15 (31.9) | 17<br>(50.0) | 0 (0) | 0 (0) | .... |
| BMI kg/m<sup>2</sup> mean<br>(%) | 29.2<br>(7.2) | 33.1 (5.7) | 29.5<br>(6.1) | 32.4<br>(8.2) | 31.9 (7.2) | 0.361* |
| BMI greater than<br>30kg/m<sup>2</sup> n (%) | 10<br>(43.4) | 31 (66.0) | 14<br>(41.2) | 8 (47.1) | 17 (56.7) | 0.177 |
| RCRI score 1/2<br>n (%) | 6<br>(26.1) | 8 (17.0) | 7 (20.6) | 3 (17.7) | 15 (48.4) | 0.024 |
| Hb g/dl mean<br>(SD) | 13.6<br>(1.7) | 14.7 (1.8) | 14.6 (<br>1.8) | 14.2 (<br>1.9) | 13.4 (2.9) | 0.081 |
| Low HB n (%) | 6<br>(26.1) | 4 (8.5) | 5 (14.7) | 3 (17.7) | 8 (25.8) | 0.204<sup>μ</sup> |
| eGFR mean (SD) | 79.9<br>(20.3) | 79.0 (18.7) | 79.9<br>(17.3) | 86.5<br>(22.1) | 86.6<br>(11.3) | 0.436* |
| GFR <60 n (%) | 4<br>(17.4) | 8 (17.0) | 2 (6.1) | 2 (12.5) | 1 (3.5) | 0.268<sup>μ</sup> |
| Diabetes mellitus<br>n (%) | 5<br>(21.7) | 6 (12.8) | 4 (11.8) | 4 (23.5) | 3 (9.7) | 0.559 |
| Hypertension<br>n (%) | 16<br>(69.6) | 29 (61.7) | 18<br>(52.9) | 10<br>(58.8) | 15 (48.4) | 0.547 |
| Other co-<br>morbidities | 5<br>(21.7) | 7 (14.9) | 8 (23.5) | 5 (29.4) | 7 (22.6) | 0.743 |

\*p-value for robust test of equal variance; \*\*p-value for K-test of equality of median; μ p-value for Fisher's exact test

## Results – Length of stay (LOS)

The median LOS for knee surgery, hip surgery, breast surgery and uterine surgery was 3.5 days, 4.0 days, 3.0 days and 3.5 days respectively (Figure 5). A total of a 152 patients (71%) stayed in hospital for more than a day or longer, 15 were admitted into an intensive care unit (ICU) and 99 into a high care unit (HCU). The median LOS was 3.5 days (IQR 2.8-4.5 days). A total of a 101 (47.2%) patients were admitted for more than 3 days. (Figure 6)

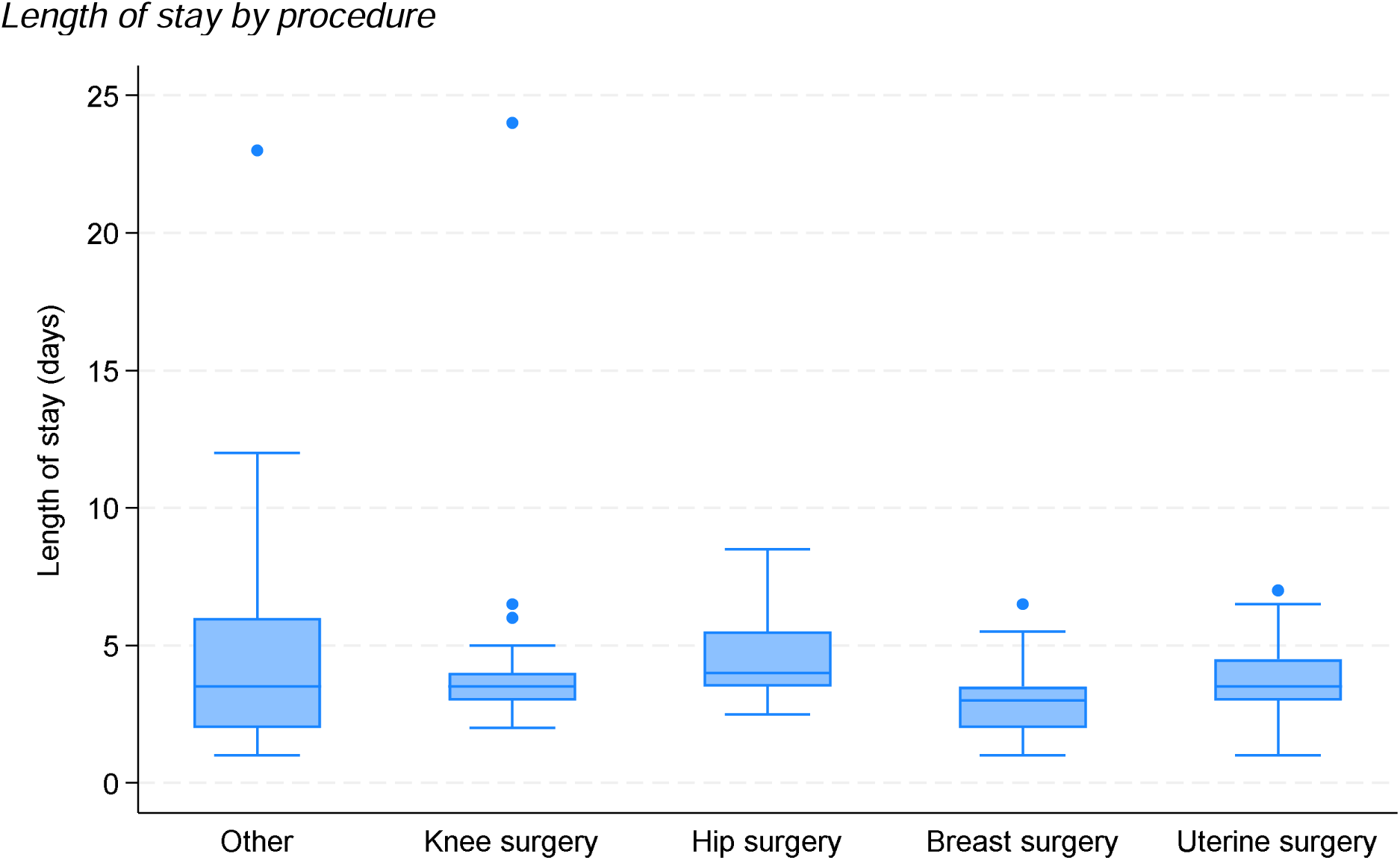
Other=3.5 days (IQR 2- 6 days) ; Knee surgery = 3.5 (IQR 3- 4) ; Hip surgery = 4.0 (IQR 3.5- 4.5); Breast surgery = 3 (IQR 2- 3.5); Uterine surgery 3.5 days (3- 4.5 days) ; p-value for equality of medians = 0.022

**Figure 6:**
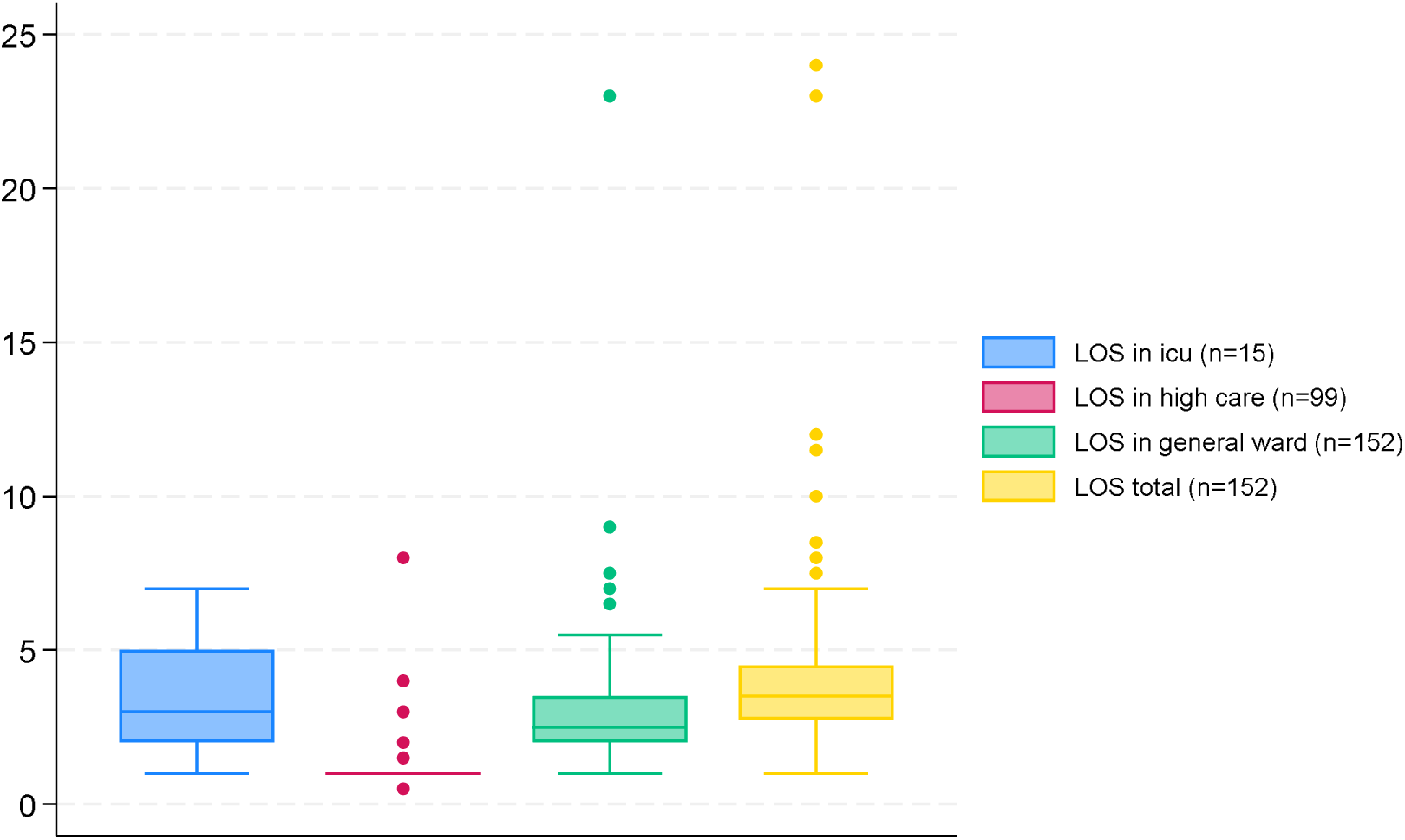
Distribution of length of hospital stay among those who stayed in hospital post-op.

The orthopaedic procedures (hip [p=0.001] and knee surgery [p=0.039]) were more likely to be admitted to ICU or High care than either breast or uterine surgical procedures. (Table 5).

**Table 5:** Factors associated with admission to ICU or high care post-surgery among patients seen in the pre-operative clinic.

| Variable | % admitted to ICU/<br>high care post-op | Univariable RR<br>(95% CI) | p-value | Multivariable<br>RR<br>(95% CI) | p-value |
| --- | --- | --- | --- | --- | --- |
| Age in years |  | 1.01 (1.00- 1.02) | 0.055 | 1.00 (0.99- 1.02) | 0.808 |
| Sex |  |  |  |  |  |
| Female | 67/162 (41.4) | 1.00 |  | 1.00 |  |
| Male | 32/52 (61.5) | 1.49 (1.12- 1.97) | 0.006 | 1.01 (0.75-1.34) | 0.970 |
| Race |  |  |  |  |  |
| Other | 16/38 (42.1) | 1.00 |  |  |  |
| White | 83/176 (47.2) | 1.12 (0.75- 1.68) | 0.584 |  |  |
| Year |  |  |  |  |  |
| 2021 | 80/154 (52.0) | 1.64 (1.10- 2.45) | 0.016 | 1.57 (1.06- 2.31) | 0.023 |
| 2022 | 19/60 (31.7) | 1.00 |  | 1.00 |  |
| *BMI* |  | 0.99 (0.97- 1.01) | 0.539 |  |  |
| *One or more comorbidities* |  |  |  |  |  |
| No | 20/42 (47.6) | 1.00 |  |  |  |
| Yes | 79/172 (45.9) | 0.96 (0.67- 1.38) | 0.843 |  |  |
| *ASA grading* |  |  |  |  |  |
| 1 | 6/12 (50.0) | 1.27 (0.61- 2.64) | 0.518 | 1.06 (0.53- 2.15) | 0.866 |
| 2 | 81/173 (46.8) | 1.19 (0.73- 1.94) | 0.481 | 1.23 (0.76- 1.99) | 0.401 |
| 3 | 11/28 (39.3) | 1.00 |  | 1.00 |  |
| *RCRI points* |  |  |  |  |  |
| 0 | 71/161 (44.1) | 1.00 |  | 1.00 |  |
| 1/2 | 28/53 (52.8) | 1.20 (0.88- 1.63) | 0.252 | 1.33 (0.99- 1.78) | 0.054 |
| *eGFR* |  | 0.99 (0.983- 0.998) | 0.017 | 1.00 (0.988- 1.00) | 0.199 |
| *Hypertension* |  |  |  |  |  |
| No | 40/88 (45.5) | 1.00 |  |  |  |
| Yes | 59/126 (46.8) | 1.03 (0.77- 1.38) | 0.844 |  |  |
| *Diabetes mellitus* |  |  |  |  |  |
| No | 83/180 (46.1) | 1.00 |  |  |  |
| Yes | 16/34 (47.1) | 1.02 (0.69- 1.51) | 0.919 |  |  |
| *Planned Procedure* |  |  |  |  |  |
| Other | 11/34 (32.4) | 1.00 |  | 1.00 |  |
| Knee surgery | 39/71 (54.9) | 1.70 (1.00- 2.89) | 0.051 | 1.74 (1.03- 2.95) | 0.039 |
| Hip surgery | 32/45 (71.1) | 2.20 (1.30- 3.70) | 0.003 | 2.38 (1.39- 4.05) | 0.001 |
| Breast | 6/21 (28.6) | 0.88 (0.38- 2.03) | 0.770 | 0.97 (0.42- 2.25) | 0.940 |
| Uterine | 11/43 (25.6) | 0.79 (0.39- 1.60) | 0.514 | 0.66 (0.31- 1.37) | 0.264 |

There was a trend towards admitted patients with a RCRI score of 1 or 2 to ICU or High care but it did not reach significance (P=0.054).

There was no association between pre-operative ASA grading and utilisation of High care or ICU post operatively (ASA 1 [p=0.866], ASA 2 [0.401]).

Females were more likely to be admitted for longer than 3 days (Table 6) as compared to their male counterparts (p=0.003).

**Table 6:**
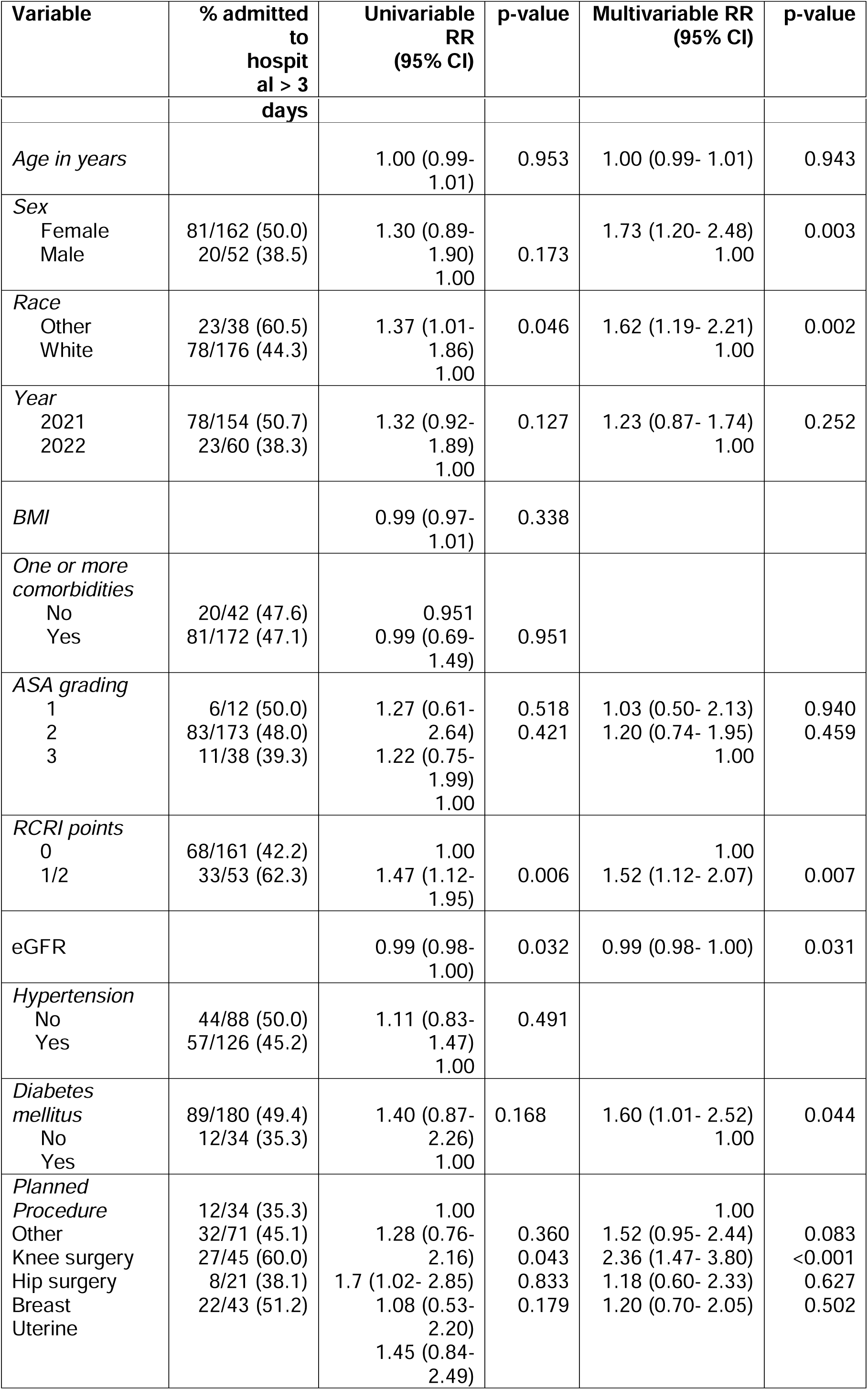
Factors associated with admission for > 3 days’ post-surgery among patients seen in the pre-operative clinic.

Patients with an RCRI score of 1 or greater were more likely to be admitted for a prolonged period (p= 0.07) as were those patients who had undergone knee surgery (p=<0.001)

Diabetic patients were less likely to have spent more than 3 days in hospital (p=0.044).

LOS per procedure type was assessed against the following risk factors : ASA grade, estimated glomerular filtration rate (eGFR), low Hb (<13.8g/dl in males ; <12.4g/dl in females), obesity (defined as BMI>=30kg/m^2^), presence of diabetes mellitus.

There was only a significant association with a low eGFR in knee surgery (p=0.027) and age in hip surgery (p=0.049) (Table 7).

**Table 7.** : Length of stay per procedure and associated possible risk factors.

*Length of stay by ASA grading and procedure*
| **ASA grading** | **Knee surgery**<br>**(median, IQR ,**<br>**n)** | **Hip surgery**<br>**(median, IQR ,**<br>**n)** | **Breast**<br>**surgery**<br>**(median, IQR ,**<br>**n)** | **Uterine**<br>**(median, IQR**<br>**, n)** |
| --- | --- | --- | --- | --- |
| 1 | 3.5 (3.5- 5.5 ),<br>3 | 4 (3.25- 4.75 ),<br>4 | --- | 5 (5- 5), 1 |
| 2 | 3.5 (3 – 4 ) , 38 | 4 (3.5 – 5.5 ) ,<br>26 | 3 (2- 3.5 ) , 15 | 3.5 (3- 4.25),<br>24 |
| 3 | 3 (2.5 – 4), 5 | 5 (3.5- 7) , 4 | 2.25 ( 1- 3.5 ) ,<br>2 | 3.5 (3.5- 4.5 ) ,<br>6 |
| p-value** | 0.317 | 0.767 | 0.929 | 0.434 |

\*\*K-test for equality of medians

*Length of stay by renal dysfunction and procedure*
| **GFR** | **Knee surgery**<br>**(median, IQR ,**<br>**n)** | **Hip surgery**<br>**(median, IQR ,**<br>**n)** | **Breast**<br>**surgery**<br>**(median, IQR ,**<br>**n)** | **Uterine**<br>**(median, IQR**<br>**, n)** |
| --- | --- | --- | --- | --- |
| GFR <60 | 4 (3.5- 6.25), 8 | 3.75 (2.5- 5), 2 | 4 (3- 5), 2 | 6.5 (6.5- 6.5),<br>1 |
| GFR ≥60 | 3.5 (3- 4), 39 | 4 (3.5- 5.5), 31 | 3 (2- 3.5), 14 | 3.5 (3- 4.25),<br>28 |
| p-value* | 0.027 | 0.640 | 0.550 | 0.138 |

\*Ranksum test

*Length of stay by anemia and procedure*
| **Hb** | **Knee surgery**<br>**(median, IQR ,**<br>**n)** | **Hip surgery**<br>**(median, IQR ,**<br>**n)** | **Breast**<br>**surgery**<br>**(median, IQR ,**<br>**n)** | **Uterine**<br>**(median, IQR**<br>**, n)** |
| --- | --- | --- | --- | --- |
| Normal Hb | 3.5 (3- 4), 43 | 3.5 (3- 4) , 29 | 2.8 (2- 3.5), 14 | 3.5 (3- 4.5), 23 |
| Low Hb | 3.8 (2.75- 4) 4 | 5.5 (5- 8), 5 | 3.5 (1- 5.5), 3 | 3.5 (2.5- 4.25), |
|  |  |  |  | 8 |
| p-value* | 0.848 | 0.103 | 0.762 | 0.662 |

\*Ranksum test; Low Hb was defined as: Hb males 13.8-18.8 g/dL, Hb females 12.4-16.7 g/dl

Length of stay by BMI and procedure
| **BMI category** | **Knee surgery**<br>**(median, IQR ,**<br>**n)** | **Hip surgery**<br>**(median, IQR ,**<br>**n)** | **Breast**<br>**surgery**<br>**(median, IQR ,**<br>**n)** | **Uterine**<br>**(median, IQR**<br>**, n)** |
| --- | --- | --- | --- | --- |
| <30 | 3.5 (2.75- 4 ),<br>16 | 4 ( 3.5 - 5.5),<br>20 | 2.5 ( 2- 3.5), 9 | 4 (3.5- 4.5 ),<br>13 |
| ≥30 | 3.5 ( 3- 4), 31 | 4 (3.5 - 5), 14 | 3.3 (1.8 - 4.3) ,<br>8 | 3.5 (2.5- 3.5),<br>17 |
| p-value* | 0.508 | 0.923 | 0.826 | 0.080 |

\*Ranksum test

Length of stay by diabetes and procedure
| **Diabetes** | **Knee surgery**<br>**(median, IQR ,**<br>**n)** | **Hip surgery**<br>**(median, IQR ,**<br>**n)** | **Breast**<br>**surgery**<br>**(median, IQR ,**<br>**n)** | **Uterine**<br>**(median, IQR**<br>**, n)** |
| --- | --- | --- | --- | --- |
| No | 3.5 (3- 4), 41 | 4 (3.5 - 5.5 ),<br>30 | 3.5 (2 - 3.5) ,13 | 3.5 (3.3 - 4.5),<br>28 |
| Yes | 3.5 (3 - 4) , 6 | 5.3 (3.5- 7) , 4 | 2.5 (1.5 - 4), 4 | 3 (1.5 - 3.5 ) ,<br>3 |
| p-value* | 0.650 | 0.336 | 0.577 | 0.148 |

\*Ranksum test

Length of stay by age category and procedure
| **Age in years** | **Knee surgery**<br>**(median, IQR ,**<br>**n)** | **Hip surgery**<br>**(median, IQR ,**<br>**n)** | **Breast**<br>**surgery**<br>**(median, IQR ,**<br>**n)** | **Uterine**<br>**(median, IQR**<br>**, n)** |
| --- | --- | --- | --- | --- |
| <50 | 3.5 ( 3 - 3.5), 5 | 3.8 ( 3 – 4), 4 | 5.5 (5.5- 5.5) ,<br>1 | 3.5 ( 2- 4), 14 |
| 50- 65 | 3.5 (2.5 - 3.75),<br>16 | 4 (3.5 - 5), 17 | 3.5 (3.5- 3.5), 5 | 3.5 (3.5 -<br>4.5), 12 |
| >65 | 4 ( 3.5- 5), 26 | 4.5 ( 3.5 – 6),<br>13 | 2.5 ( 2 - 3.5),<br>11 | 3.5 ( 3- 6), 5 |
| p-value** | 0.111 | 0.049 | 0.081 | 0.951 |

\*\*K-test for equality of medians

## Discussion

The association between length of stay and an increased RCRI score in our population is in keeping with previous studies in which it was noted that there was an increase in length of stay in patients with an RCRI score of 3 and greater ^(10)^ ^(19)^ ^(20)^. This could be due to the fact that these patients require closer monitoring due to their increased risk factors. We did not show a significant increase in ICU or High care usage according to the RCRI in our sample however there was a signal that trended towards significance. Patients with increased risk factors may also have delayed recovery and rehabilitation may be a more gradual process as healthcare practitioners may show increase caution in dealing with these particular patients.

In our study ASA grading was not associated with an overall prolonged length of stay. This is not in keeping with findings from other studies in which there was an increase in length of stage between ASA grade 2 and Grade 3^(21)^.

Furthermore, the need for admission to ICU post op was not influenced by the patients pre-operative ASA grading. This was also not in keeping with other studies in which patients with higher pre-operative ASA gradings spent a longer period in ICU post op ^(22)^.

A possible explanation for the above findings is that elective surgeries in private tend to be largely protocolised. This results in patients being admitted to ICU or high care regardless of their risk profile and this is an area that will need to be altered based on future guidelines.

Approximately 25.8% of patients booked for elective uterine surgical procedures were found to be anaemic. This was greater than patients undergoing elective knee (8.5%), hip (14.7%) or breast surgery (17.7%). This is unsurprising as patients undergoing uterine surgery are more likely to have experienced some form of abnormal uterine bleeding resulting in anaemia ^(23)^. It is somewhat surprising that anaemia did not result in an extended length of stay. A study by Wang et al ^(17)^ in patients undergoing non-cardiac and non-obstetric surgery showed that patients who stayed longer than 7 days had, on average, a lower preop Hb compared to those patients who stayed 7 days or less. Furthermore, a preoperative Hb of less than 11.9g/dl had a decrease in LOS by 2 days for every 1g/dl increase in Hb. This was again noted in a study performed by Bulte et al ^(24)^ which highlighted the association of anaemia on LOS with an increase on average of 1.3 days noted in patients with moderate to severe anaemia. A systemic review and meta-analysis by Fowler et al ^(25)^ indicated that pre-operative anaemia was associated with increased risk of mortality (Odds Ratio [OR] 2.9), infection (OR 1.93) and acute kidney injury (OR 3.75). All of the above could result in a prolonged LOS. A possible explanation for why we did not show an increased LOS in patients with a pre-op anaemia is that once identified patients were provided with iron supplementation and advised to discontinue drugs that may worsen iron deficiency (eg. Anti-inflammatories). Minimally invasive surgical techniques may also have prevented significant blood losses in theatre. A form of selection bias in that only patients who were considered fit for surgery and did not require significant optimisation moved forward to having their planned procedures. This resulted in fewer complications and thus no impact on length of stay.

Chronic renal dysfunction noted has an impact on morbidity and mortality in patients undergoing surgery ^(26)^. In particular, Liao et al showed that patients with renal insufficiency (GFR<60ml/min) were hospitalised for a longer period than those without renal insufficiency. (3 days vs 2 days; P value <0.0001) ^(18)^

In our study, patients undergoing elective knee surgery with pre-operative eGFRs of <60ml/min were noted to have an increased length of stay which is in keeping with the literature. There were very few patients undergoing other surgeries who had a decreased eGFR pre-op which may have been due to the smaller sample sizes in those groups.

Obesity is a major cause of morbidity and may result in a reduced life expectancy^(27)^. It is estimated that currently there are over a billion people worldwide classified as obese (BMI>=30) ^(28)^. A large meta-analysis by Plassmeier et al. ^(29)^ indicated that patients who were obese had longer hospital stays after undergoing colorectal surgery, upper gastrointestinal procedures and pancreatic surgery. Patients who underwent total hip arthroplasty were also noted to have had an increased LOS ^(30)^. Obesity did not result in an extended length of stay in our study. This is in keeping with other studies in which LOS was not impacted by obesity^(31)^. In particular, obesity was not associated with an extended length of stay following primary hip arthroplasty in a South African setting^(32)^. A possible explanation for this is that our study population consisted of patients in a private healthcare setting. These patients generally have increased access to healthcare resources ensuring that co-morbidities are detected and managed earlier. This should lower their overall cardiometabolic risk and decrease subsequent complications post op which may have resulted in no increase in LOS.

Type 2 Diabetes was a co-morbidity in 30 patients (14%). There are multiple studies which report an increase in length of stay in diabetic patients compared to non-diabetic patients ^(33–36)^. There was no increase in admission to high care or ICU, or an increase in LOS noted in patients with type 2 diabetes in our cohort. This is in keeping with a previous study by Lejeune et al in which type 2 DM did not confer a risk for increase LOS in patients undergoing colo-rectal surgery ^(37)^. This was attributed to the multi-disciplinary approach to patient care in the peri-operative period. The patients in our cohort were aware of their type 2 diabetic status and were educated in the pre-op clinic to ensure tight control in the peri-operative period. The increased awareness around the patients Type 2 diabetes status by a multi-disciplinary team may, in our case, also have limited the deleterious effects often experienced by these patients in the post operative period such as post operative pneumonia ^(35)^ and urinary tract infections ^(38)^.

Age has been identified as a risk factor in predicting an extended LOS post operatively ^(13, 39, 40)^. Ageing is associated with frailty and disability ^(41, 42)^ which will decrease a patients mobility and the ability to withstand a severe stressor such as invasive surgery. Dlamini et al also showed in their study that maximum walking distance of less than a 100m resulted in an extended length of stay ^(32)^ and the older, frail patient may fall in this category. A previous study looking at outcomes post hip and knee arthroplasty in a New Zealand population noted the LOS generally increased with age except in the age group less than 40 years ^(43)^.

In our study, patients over the age of 65 years who had undergone hip surgery had a prolonged LOS (4.5 days) compared to patients between 50-65yrs (4 days) and patients under 50 years (3.8 days) [p value 0.049]. There was no difference in LOS based on age in the other 3 main surgical procedures (knee, uterine, breast).

The average length of stay in patients undergoing hip arthroplasty was longer than patients undergoing knee arthroplasty in each of the age ranges (<50 yrs; 50-65yrs; >65 yrs). There was no significant difference in age between the patients in our cohort who had undergone knee and hip arthroplasty (63.7yrs vs 63.2 yrs respectively). In the previously mentioned New Zealand study ^(43)^ 29.9% of patients undergoing a primary hip replacement had a LOS longer than 5 days compared to 34.2% of patients who had undergone a primary knee replacement.

A possible reason for our findings may be procedural in nature. An anterior approach compared to a posterior approach in hip replacements may result in shorter LOS ^(44)^. However, our study did not include any information regarding type of procedure which would have allowed us to validate this theory.

This study provides valuable insights into the demographic and clinical characteristics of patients referred to a preoperative clinic in a private practice in Johannesburg and highlights the factors influencing postoperative length of stay.

This study has several limitations that must be acknowledged. Firstly, the study was conducted in a single private preoperative clinic in Johannesburg, which may limit the generalisability of the findings to other settings, particularly public hospitals or clinics in other regions of South Africa where patient profiles and resource availability may differ.

Secondly, the relatively small sample size, particularly within subgroups such as patients with renal dysfunction or undergoing specific types of surgeries, may limit the statistical power to detect significant associations between certain risk factors and length of stay. A larger cohort may have provided more robust results, especially in assessing the impact of variables like ASA grading and RCRI on postoperative outcomes.

Thirdly, the study did not account for the potential influence of surgical techniques, such as anterior versus posterior approaches in hip replacement surgeries, which may affect the length of stay. Future studies incorporating procedural differences may provide a clearer understanding of their role in recovery times.

Additionally, the study relied on medical records for data collection, which could be subject to information bias if some patient data were incomplete or inaccurately recorded. The potential for confounding variables, such as variations in postoperative care protocols or patient compliance with preoperative instructions, was also not fully explored, which may influence the findings.

Finally, this study did not include long-term postoperative outcomes, such as readmission rates or complications after discharge, which could provide a more comprehensive view of the factors affecting recovery and healthcare utilization.

By acknowledging these limitations, future research can focus on addressing these gaps to further refine perioperative care and improve patient outcomes in a South African setting.

## Data Availability

https://doi.org/10.5061/dryad.sbcc2frh1

